# Birth anthropometry among three Asian ethnic groups in Singapore – new growth charts

**DOI:** 10.1101/2022.03.30.22273158

**Authors:** Sonoko Sensaki, Yinan Mao, Agnihotri Biswas, Chinnadurai Amutha, Zubair Amin, Alex R Cook, Jiun Lee

## Abstract

**Objective:** We analyse birth anthropometry of Asian babies and its socioeconomic exposures, develop gestational age and gender-specific birth anthropometry charts and compare to the widely used Fenton chart.

**Design:** Retrospective observational study.

**Setting:** Department of Neonatology at the National University Hospital in Singapore.

**Population or sample:** We report data from 52 220 Chinese, Indian and Malay infants, born from 1991-1997 and from 2010-2017 in Singapore.

**Methods:** The BW, length and head circumference are each modelled with maternal exposures using general additive model. Anthropometry charts are built using smoothed centile curve and compared with Fenton charts using binomial test.

**Main outcome measures:** BW, head circumference, crown-heel length.

**Results:** In contrast to the marked differences in birth anthropometry among these ethnic populations, when exposed to a uniform socioeconomic environment, their intrauterine growth and birth anthropometry were almost identical. From the gestational age specific anthropometric charts, until about late prematurity, Asian growth curves, as derived from our cohort, mirrored that of Fenton’s; thereafter, Asian babies showed a marked reduction in growth velocity.

**Conclusions:** These findings suggest comparative slowing of intrauterine growth among Asian babies towards term gestation. This phenomenon may be explained by two possible postulations, firstly, restrictive effects of a smaller uterus of shorter Asian women towards term and secondly, early maturation and senescence of fetoplacental unit among Asians. In clinical practice the new birth anthropometry charts will more accurately identify true fetal growth restriction as well as true postnatal growth failure in preterm infants when applied to the appropriate population.

**Funding:** Singapore Population Health Improvement Centre (NMRC/CG/C026/2017_NUHS).

## Introduction

Birth anthropometry, especially birth weight (BW), is an important determinant of childhood and future adult health ^1–11^. The developmental origins of health and disease theory posits that neonates with lower BW are at greater risk of perinatal mortality ^12^ and chronic conditions in later life such as type-2 diabetes ^1,2^, obesity ^3–5^, cardiovascular diseases ^6^, hypertension ^7,8^, certain cancers ^9^, poor neurocognitive development ^10,11^, and mental disorders ^13^. Significant differences in BW and low BW incidence have been found among different countries and ethnicities ^14–16^. Although such differences may be the result of modifiable exposures such as maternal nutrition ^17^, perinatal care ^18^, socioeconomic disparities ^18^, or smoking ^19,20^, some of the variability may have its origins in genetic differences ^21^. Because standard growth charts were developed in populations in which European ancestry predominates, if these references are used as norms for babies with non-European ancestry, it is possible that babies of non-European ancestry that are of normal size for their ethnic heritage may erroneously be classified as small for gestational age, or as having microcephaly.

The three main population centers of Asia—East, South East, and South—which as a group contain about half of the world’s population, display striking differences in BW: neonates weigh 3200g in China on average, 2900g in Indonesia, and between 2600g and 2800g in India ^14^. Singapore, a city state off the southern tip of the Malay peninsula, is a microcosm of Asia as a whole, with a Chinese majority and large minorities of Indians and Malays. Although there remain socio-economic ^22^ and health ^23^ disparities between Singaporeans of these three ethnic groups, these differences are much less pronounced than between people in territorial China, India and Singapore’s neighbors in the Malay Archipelago. Singaporean birth cohorts thus provide two unique opportunities: firstly, a controlled opportunity to quantify differences in birth anthropometry between East, South East and South Asians, and, secondly, given the high per capita income and excellent health outcomes in Singapore ^24^, to compare norms of birth anthropometry among Asians to international, i.e. European, levels.

To this end, we investigated the epidemiology of birth anthropometry of all 52 220 infants born at Singapore’s National University Hospital over the period 1991-1997 and 2010-2017, with the twin objectives of investigating determinants of birth anthropometry and investigating differences between local and international growth charts.

## Methods

### Population

We extracted information from a database of a birth screening program originally intended to investigate birth defects ^25^. All infants (n = 52 220) born in 1991-1997 and 2010-2017 at National University Hospital, Singapore were included. The institutional review board (National Healthcare Group Domain Specific Review Board) approved waiver of informed consent for the current study as this was a retrospective study using anonymised data (NHG DSRB Ref: 2018/00389).

### Birth anthropometry

All birth anthropometry measurements were recorded by trained hospital staff. BW was measured using calibrated digital weighing scales accurate to the nearest gram. Head circumference (HC) was measured by using the occipitofrontal circumference with a non-stretchable measuring tape. Length was measured from the top of the head to the soles of the feet using a stadiometer to the nearest centimetre. Gestational age was determined by early ultrasound dating or by last menstrual period. Gender-specific birth anthropometry charts were constructed for the cohort.

### Determinants of fetal growth

Trained interviewers collected data using structured questionnaires. The data fields included household income, maternal education, existing maternal diabetes mellitus (DM), smoking, alcohol consumption, and coffee intake. Maternal height, blood pressure, and hemoglobin value were collected from clinical record at delivery. Hypertension was determined as having a blood pressure greater than 140/90 mmHg. Anaemia was defined as haemoglobin level less than 11 g/dL. Variables for determinants of fetal growth that were available for both cohorts were gestational age, gender, maternal ethnicity, number of births, birth order and maternal diabetes. Data on maternal age, height, hypertension, anemia, education, duration of marriage, household income, smoking, alcohol and coffee intake were available only for 1991-1997 cohort.

### Statistical analysis

Possible erroneous data entry, measurement, or recording of BW, head circumference and crown-heel length were excluded from analysis by z score fences with five standard deviations, after excluding non-positive entries. Based on general medical experience, we cleaned up covariates outside chosen boundaries by replacing the values with NA to avoid impact of outliers in analysis: gestational age (<23 or > 42 weeks), maternal age (<10 years), maternal height (<138 or >200 cm), systolic blood pressure (< 70 or > 250 mm Hg), diastolic blood pressure (<40 or > 180 mm Hg), haemoglobin level (0 or > 40 g/dL).

All statistical analyses were carried out by a trained statistician (one of the authors). For each response variable (BW, head circumference, crown-heel length), we first fitted general linear regression model for single covariates (sex of child, maternal race, number of births, birth order, gestational DM, hypertension, anaemia, household income, maternal education, smoking, alcohol, daily coffee consumption, coffee consumption during pregnancy, maternal age, and maternal height), and then compared that with a general additive model with combined covariates, including gestational age.

In the image of Fenton 2013 growth charts ^26^, we created growth charts using generalized additive models for location, scale and shape^27^ using the gamlss package in R ^28^. We used natural cubic splines on gestational age and assumed a Gaussian distributional family to produce smoothed estimates of the centile curves at the 3^rd^, 10^th^, 50^th^, 90^th^ and 97^th^ percentiles; this was repeated for different demographic strata (sex and ethnicity). Reference centile curves for each week of gestational age were derived from the cpeg-gcep website implementation of the Fenton chart ^29^. We tested differences in the aforementioned centiles of BW, head circumference and birth length at each gestational week between the two cohorts. Specifically, the measurement corresponding to a given Fenton centile was used to classify the Singapore data as above or below target, and the Fenton centile used as the hypothesized proportion below target for the Singapore data in a binomial test.

## Results

Baseline characteristics are presented in Table 1. Participants in combined cohort included 25 017 female (48%) and 27 203 male (52%) births, which is in accordance with the elevated sex ratio at birth in Singapore ^30^. The ethnic breakdown consisted of 22 248 Chinese (43%), 16 006 Malay (31%), 8 543 Indian (16%) and 5 423 of other races (10%). Based on either the 1991-1997 cohort or combined cohort, Figure 1a presents mean BW by demographics, parity, maternal comorbidities, and risk factors [analogies for length (Figure 1b) and HC (Figure 1c)]. In the combined cohort, mean BW was 3103g (95% CI: 3096, 3109), 3075g (95% CI: 3067, 3083) and 3052g (95% CI: 3041, 3062), for Chinese, Malays and Indians, respectively. Maternal education and household income were associated with birth anthropometry in a gradient-dependent manner. Babies born to mothers with university education were likely to be heavier by 109g (95% CI 89, 130) as compared to mothers with primary education. Similarly, there was a difference of 91g (95% CI 74, 108) in mean BW between children whose households had an income more than $3000 as compared to ones below $1500. In univariate analysis, maternal ethnicity, birth order, household income, maternal education, age, height, DM, hypertension, anaemia, smoking, alcohol, were all associated with BW, HC, and length (Figure 1).

**Table 1:** Basic characteristics of the study population (1991-1997 cohort n = 21 897, combined cohort n = 52 220). Gestational age is defined as follows: Extremely preterm: to 27^6^/_7_ weeks, Very preterm: 28^0^/_7_ to 31^6^/_7_ weeks, Moderately preterm: 32^0^/_7_ to 33^6^/_7_ weeks, Late preterm: 34^0^/_7_ to 36^6^/_7_ weeks, Early term: 37^0^/_7_ weeks to 38^6^/_7_ weeks, Full term: 39^0^/_7_ weeks to 40^6^/_7_ weeks, Late term: 41^0^/_7_ weeks to 41^6^/_7_ weeks, Post term: 42^0^/_7_ weeks and beyond.

|  | 1991-1997 cohort |  | Combined cohort |  |
| --- | --- | --- | --- | --- |
|  | Count | Percent | Count | Percent |
| **Sex of child** |  |  |  |  |
| Female | 10319 | 47% | 25017 | 48% |
| Male | 11578 | 53% | 27203 | 52% |
| **Maternal race** |  |  |  |  |
| Chinese | 10485 | 48% | 22248 | 43% |
| Indian | 2811 | 13% | 8543 | 16% |
| Malay | 7621 | 35% | 16006 | 31% |
| Others | 980 | 4% | 5423 | 10% |
| **Gestational term** |  |  |  |  |
| Extremely preterm | 31 | 0% | 122 | 0% |
| Very preterm | 166 | 1% | 428 | 1% |
| Moderately preterm | 214 | 1% | 475 | 1% |
| Late preterm | 1368 | 6% | 3506 | 7% |
| Early term | 6523 | 30% | 18593 | 36% |
| Full term | 10931 | 50% | 25040 | 48% |
| Late term | 2128 | 10% | 3393 | 6% |
| Post-term | 536 | 2% | 663 | 1% |
| **Maternal age group** |  |  |  |  |
| <26 | 4216 | 19% |  |  |
| 26~30 | 8486 | 39% |  |  |
| 31~35 | 6729 | 31% |  |  |
| >35 | 2466 | 11% |  |  |
| **Maternal height group** |  |  |  |  |
| <151 | 3555 | 16% |  |  |
| 151~155 | 6606 | 30% |  |  |
| 156~160 | 7395 | 34% |  |  |
| >160 | 4341 | 20% |  |  |
| **Household income** |  |  |  |  |
| <\$1500 | 6092 | 28% | | |
| \$1500~\$3000 | 7170 | 33% | | |
| >\$3000 | 8635 | 39% | | |

**Figure 1:**
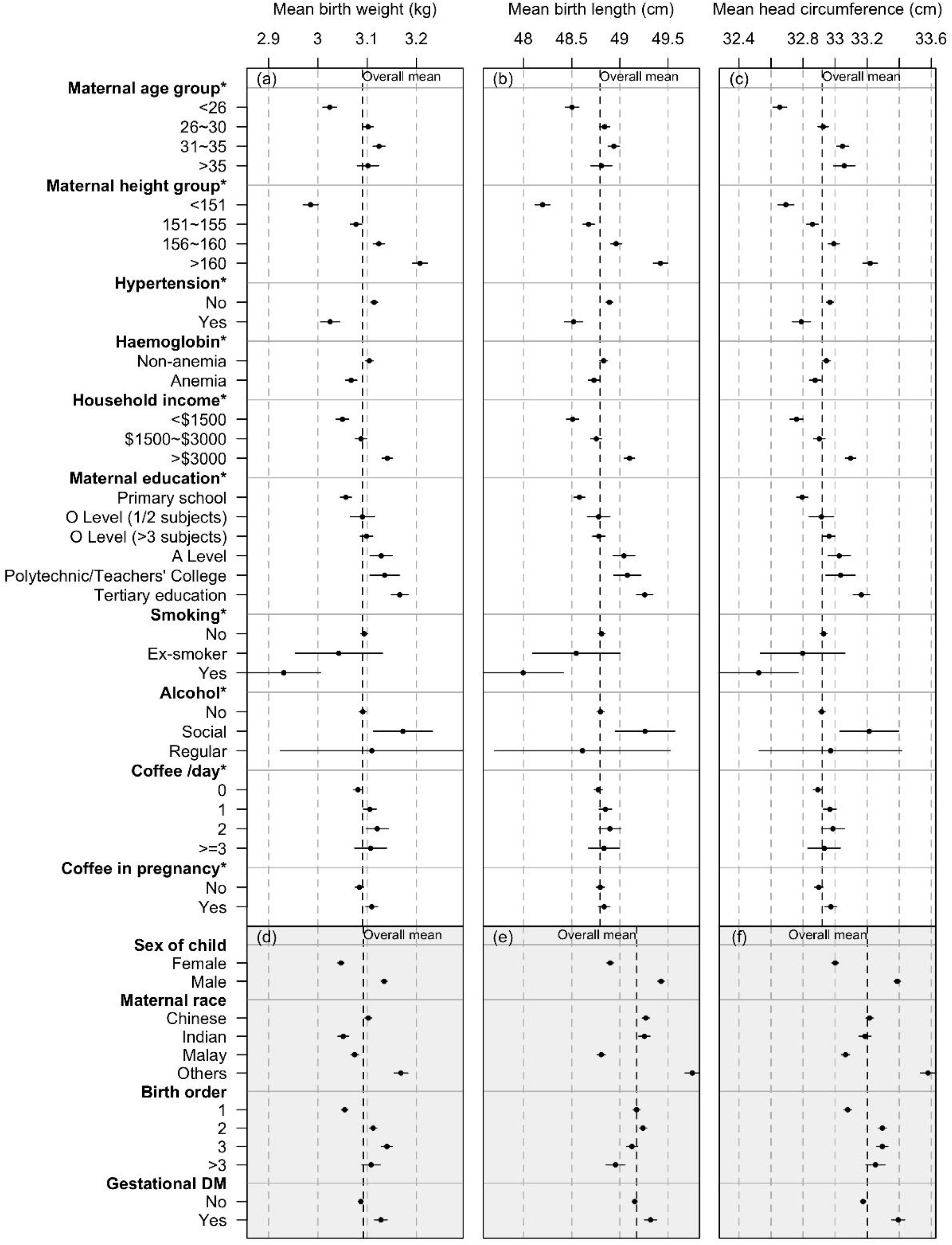
Univariate analysis of the association between BW (a), length (b) and head circumference (c) and determinants in Singapore. Lines are 95% confidence intervals. (cohort 1990-1997 n=21 897 for subplots a, b and c; combined cohort n=52 220 for shaded subplots d, e and f)

Table 2 shows the results of univariate and multivariate analysis for the association between each determinant factor and birth measurements. Based on combined cohort and after adjustment for confounders, Malay and Indian babies were slightly lighter than Chinese (by - 22g, 95% CI: -30, -14 for Malay, by -33g, 95% CI: -42, -23 for Indian). Boys were heavier than girls (by 107g, 95% CI 100, 113). Multiple births had lower BW than singletons (−320g, 95% CI: −343, −298). Increasing birth order was associated with higher BW. Babies in 2010-2017 cohort were heavier than 1991-1997 cohort (by 12g, 95%CI: 5, 19). Mothers with gestational DM have heavier babies (by 97g, 95% CI: 86, 107), but hypertension and anaemia were not significantly associated with BW after adjustment. Mothers who smoked during pregnancy had infants with lower BW than those who never smoked (by -116g; 95% CI: -180, -51). There was no significant difference in BW between mothers who were ex- or non-smokers. Mothers with alcohol and coffee intake did not show significant differences in their children’s BW. BW increased by 135g (95% CI:125,145) for every additional 10cm in maternal height. Similar to BW, our cohort’s length and HC had statistically significant association with ethnicity, sex, number of births, birth order, DM, household income, maternal education, height, and smoking (Table 2).

**Table 2:**
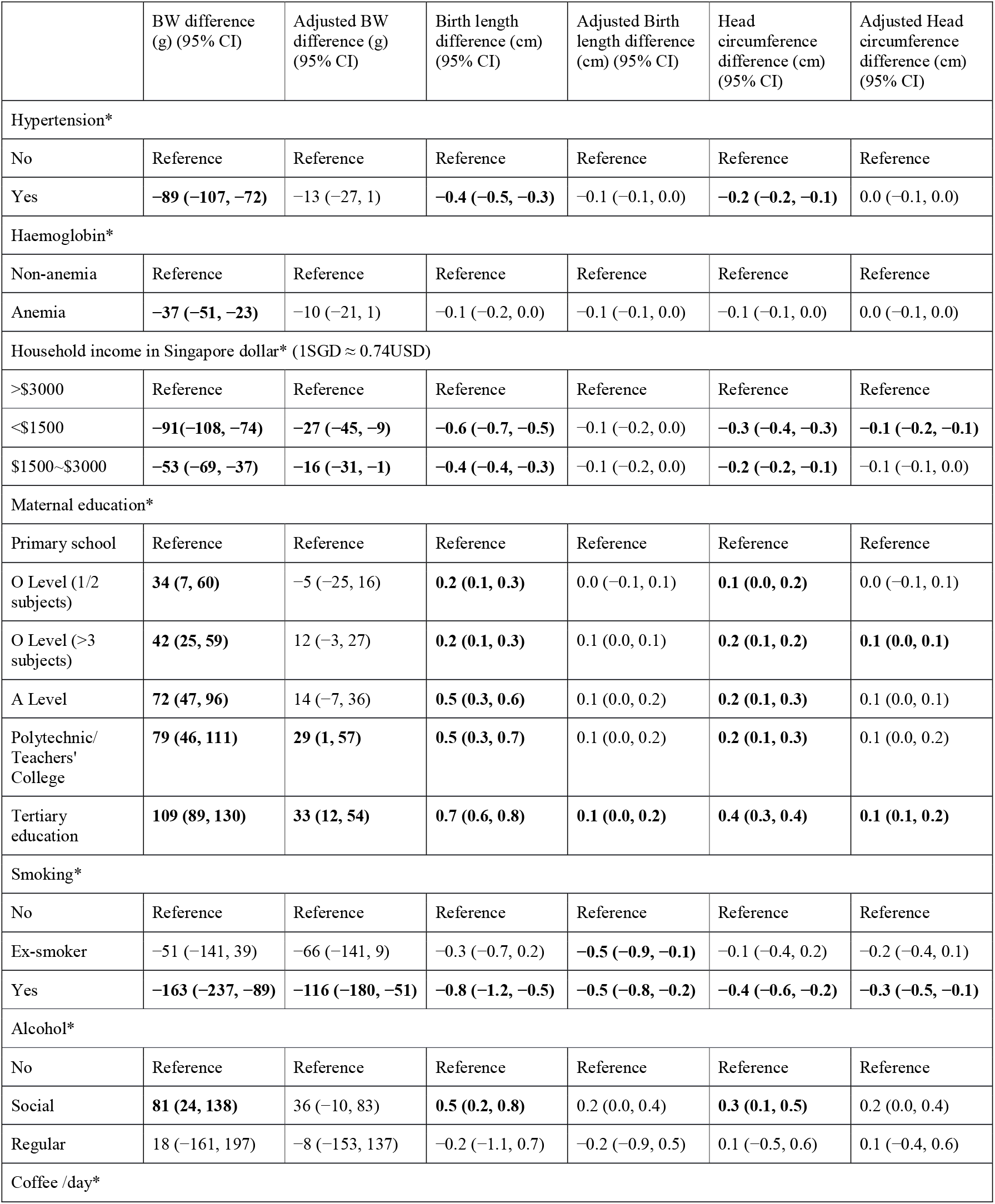
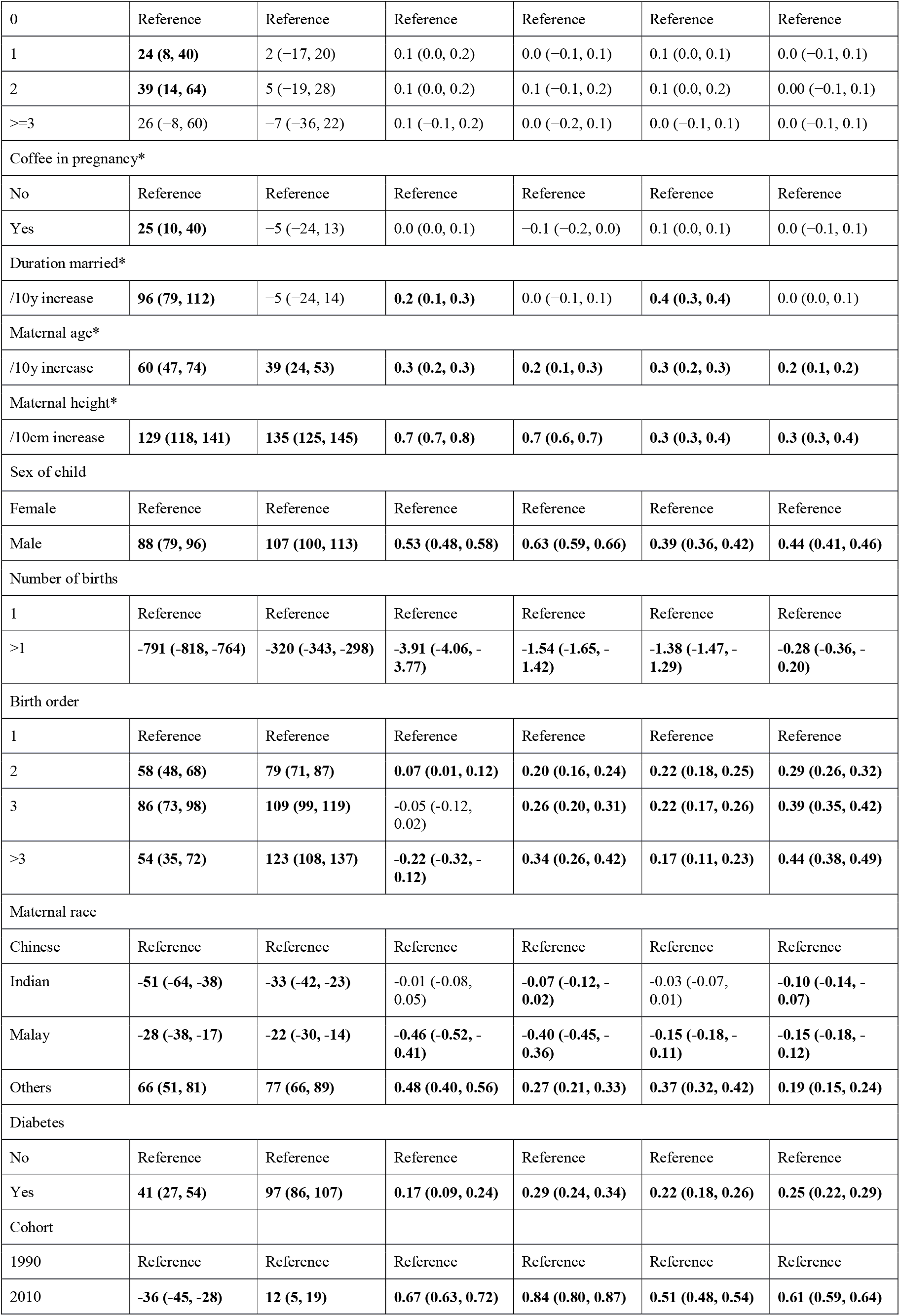
Multivariate analysis of the association between birth anthropometry and determinant factors in Singapore (1991-1997 cohort n = 21 897 in the upper part of table and marked with *, combined cohort n = 52 220 in the lower part of table). Adjusted models account for gestational age using multivariate general additive model. In 1991-1997 cohort, model is adjusted for sex of child, number of births, birth order, maternal race, and diabetes, the effects of which are omitted. CI: confidential interval. Differences significant at *α* = 0.05 are highlighted in bold.

Growth charts for BW, length and HC from the combined cohort constructed using quantile regression are presented in Figure 2 alongside the comparable quantiles from the Fenton study. There was a striking similarity in the distributions of the three anthropometrics among ours and Fenton’s data up to gestational age of 37 weeks. However, after 37 weeks, these trajectories diverged markedly, with statistically significant differences between all five reference quantiles (3^rd^, 10^th^, median, 90^th^, 97^th^). These deviations were more pronounced in the higher reference quantiles (97^th^ and 90^th^) as compared to lower ones (10^th^ and 3^rd^). These deviations cannot be solely explained by the fact that Fenton charts were likely smoothened beyond the term gestation to match postnatal growth. In particular, the growth velocity of babies in our cohort decelerates starting around 36 weeks gestation as they approach term compared to the analogous Fenton data. Until about late prematurity growth curves from our cohort mirror that of Fenton’s signifying similar rate and magnitude of intrauterine growth between European and Asian babies.

**Figure 2:**
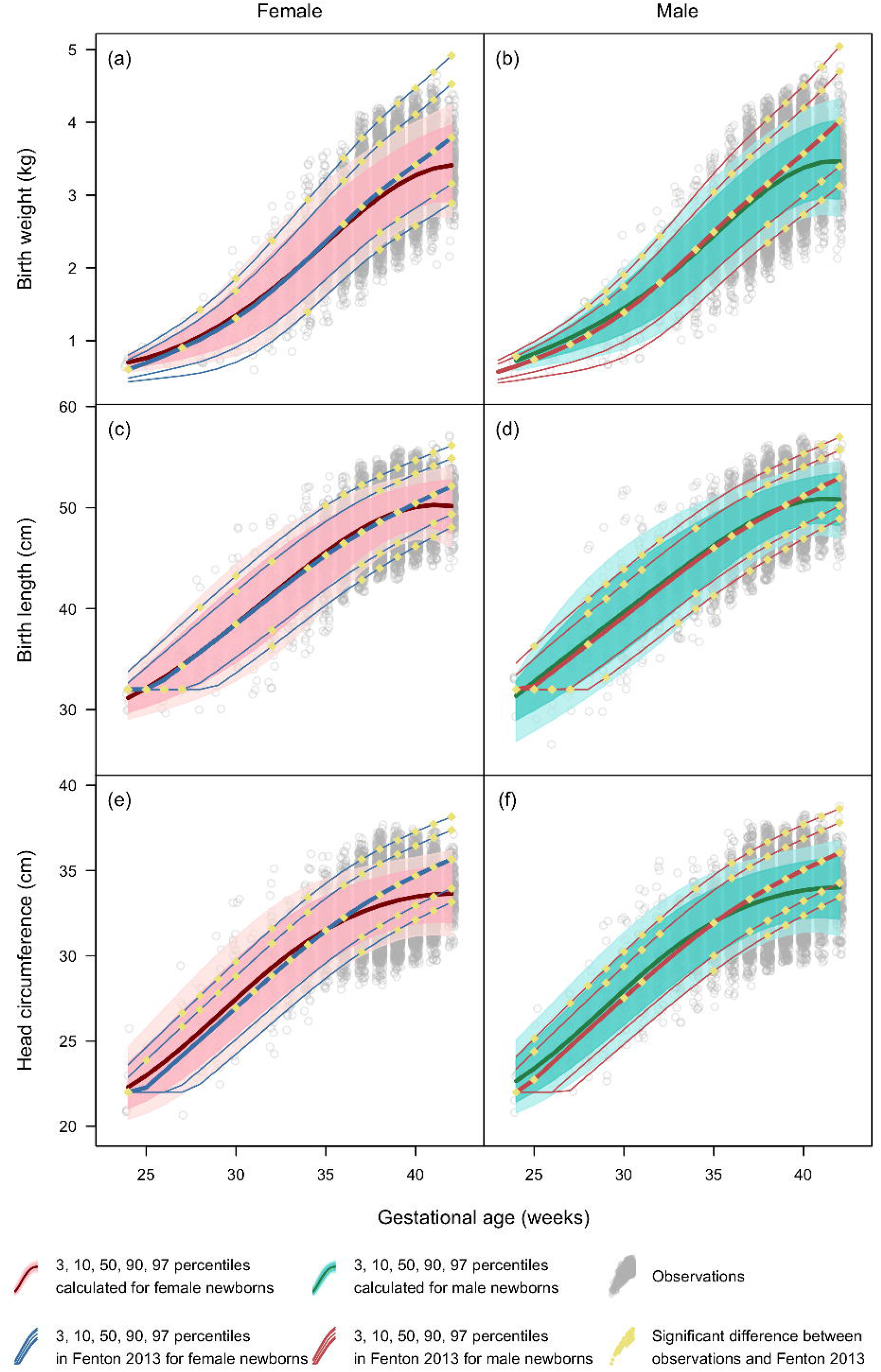
Smoothened centile charts of BW, birth length and head circumference for boys and girls by gestational age in weeks with comparison to international standard. Pink and blue shades for girls and boys show the 5 centiles calculated based on observed data points in grey, compared with baseline curves from Fenton 2013 (blue and red for female and male babies respectively). Differences in centile positions between Singapore cohort and Fenton 2013 are tested by binomial test in each gestational week separately, with significant differences marked in yellow.

**Figure 3a:**
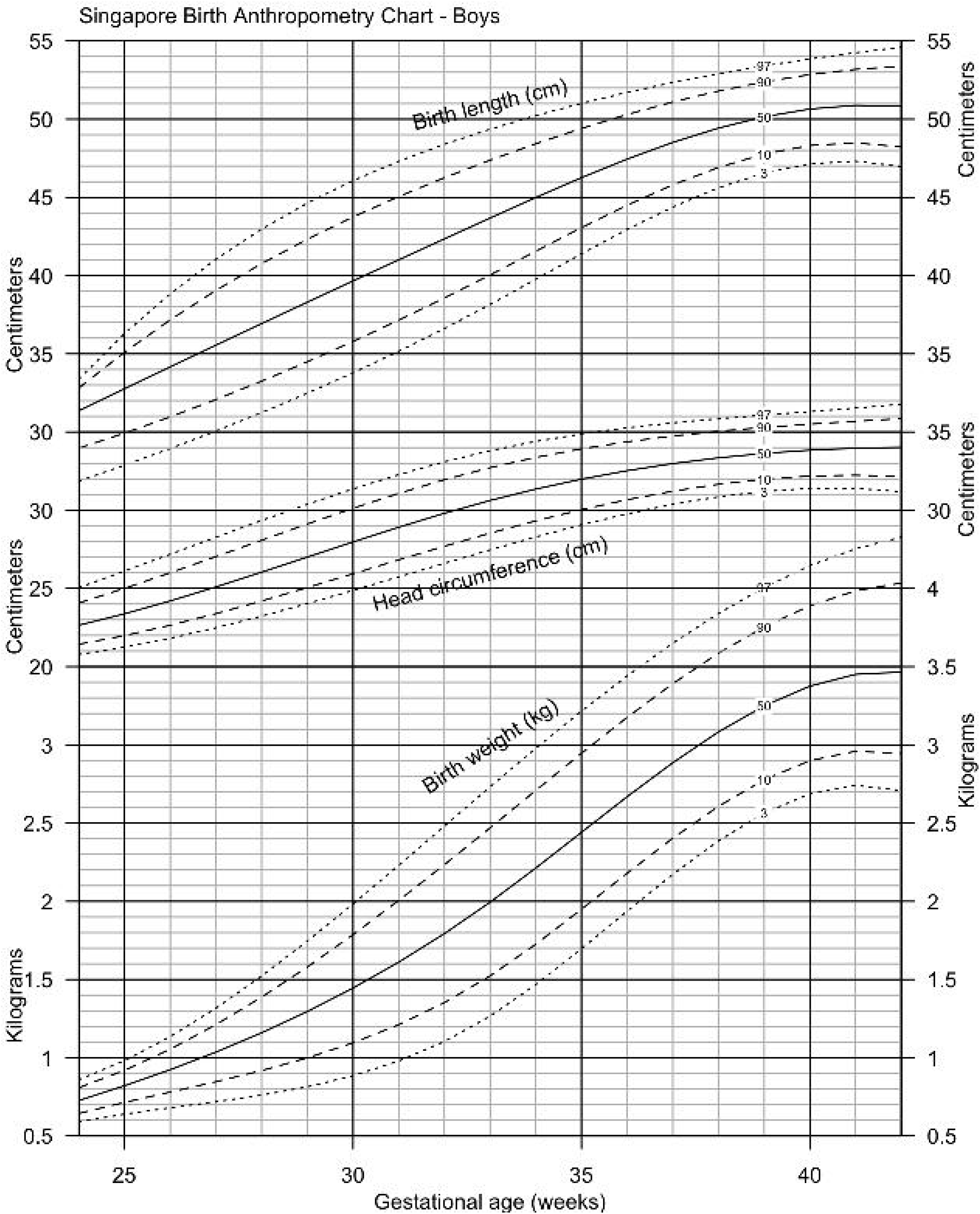
Singapore Birth Anthropometry Charts for boys. The 5 centiles curves are extracted from Figure 2 (b), (d), (f), presented in a similar fashion as Fenton 2013 for clinical reference.

**Figure 3b:**
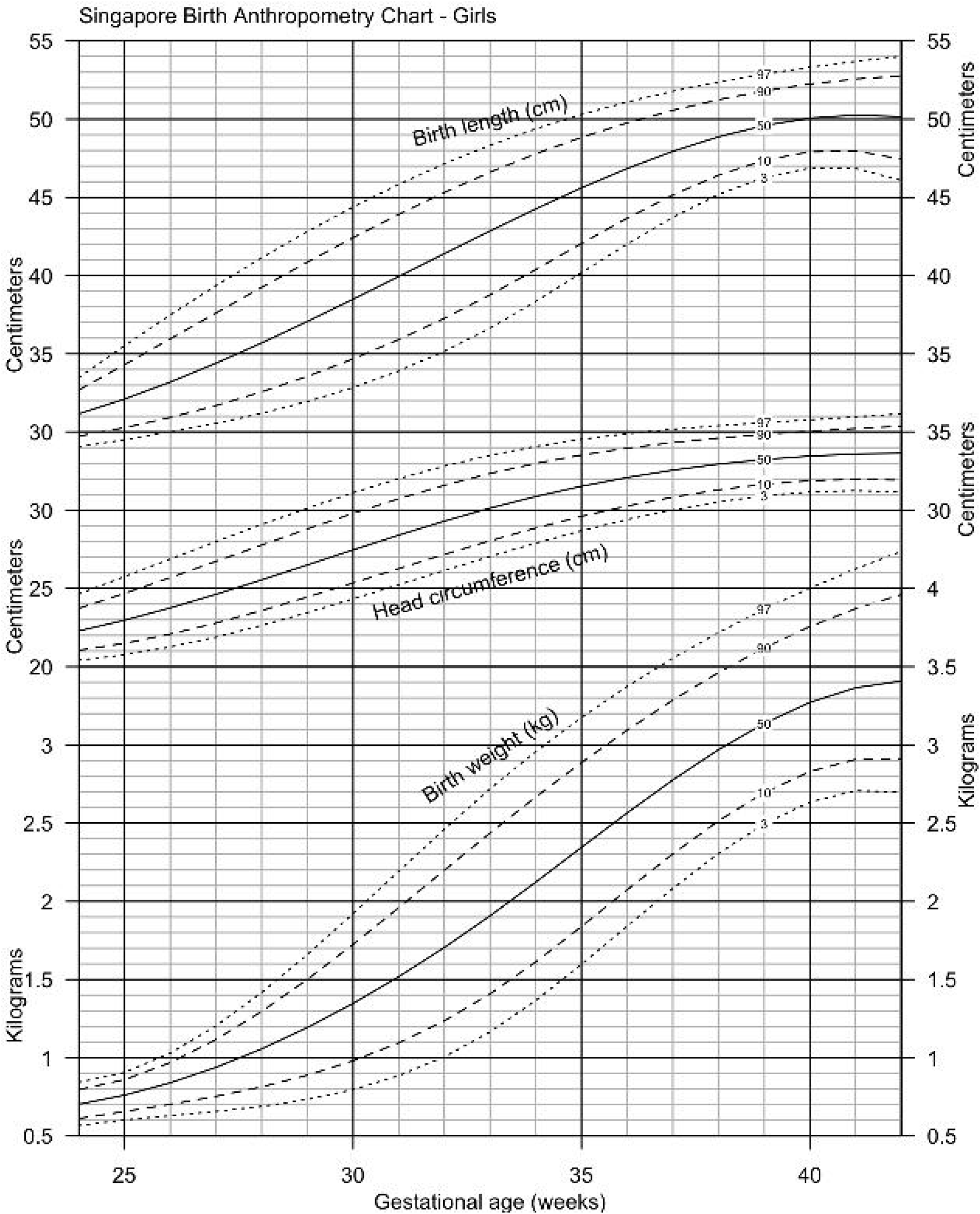
Singapore Birth Anthropometry Charts for girls. The 5 centiles curves are extracted from Figure 2 (a), (c), (e), presented in a similar fashion as Fenton 2013 for clinical reference.

## Discussion

### Main Findings

Our study presents the relationship between birth anthropometry and its contributing factors among three ethnic groups in Singapore using birth cohort data from 1991-1997 and 2010-2017. It is important to establish ethnic differences in birth anthropometry because these parameters are often used to judge the status of maternal and child health in the community.

Racial differences in BW, incidence of low BW babies and prematurity rates have been demonstrated in the United States^31^. Although this particular study argued against the influence of genetic factors governing lower BW among American-born blacks compared to African-born blacks and believed social and maternal health factors were responsible for the observed differences, a large cohort of over 10 million births demonstrated that BW and neonatal mortality among non-Hispanic whites and Hispanics were similar but non-Hispanic blacks had significantly lower BW and higher neonatal mortality^32^. This effect cannot be explained by socioeconomic and environmental influence alone. In the Asian context, a WHO study ^14^ showed marked difference in BW among ethnic Chinese, Indian and Indonesian populations residing in their country of origin. Our findings concur with the claim, as the differences in BW between Singapore’s three main ethnicities in the combined cohort were significant (though of a small magnitude), with Indians and Malays being lighter than Chinese (by −33g, 95% CI: −42, −23, by -22, 95% CI: -30, -14 respectively) after controlling for important confounders (sex of child, number of births, birth order, diabetes). Interestingly, the difference among ethnicities was not apparent in 1991 – 1997 cohort, with Indians showing marginally lower BW than Chinese (−27g, 95% CI: −48, −6) and there was no difference between Chinese and Malays before adjustment. After controlling for important confounders, there were no significant difference between Chinese and Indians, and only small difference (+40g, 95% CI: 26, 53) between Malays and Chinese. As a result, one may postulate that the large reported difference in birth anthropometry between these three Asian ethnicities in their country of origin ^14^ were due to environmental factors including socioeconomic status (SES) and perinatal care rather than genetic factors.

Consistent with previous research ^18,33–37^, we found BW to be associated with maternal height, household income, maternal education, smoking during pregnancy, existing DM and parity. These remained significant after adjustment in multivariate models. After adjusting for confounders, household income and maternal education were independently associated with birth anthropometry even in a high-income country like Singapore (1991 per capita GDP of ∼USD15 000 ranked Singapore 24^th^ highest in the world ^38^). Other SES-related factors which were not studied here such as maternal nutrition, psychosocial health, workload, and perinatal care might also mediate the relationship between SES and birth anthropometry.

### Interpretation

BW at term in our population was substantially lower than international standards (e.g., female infant at 40 weeks with a BW at 50^th^ percentile: Fenton 3415g, Singapore 3220g) despite Singapore’s generally excellent maternal and child health indicators and socioeconomic standing. Our growth curves for the distribution of birth anthropometrics (Figure 2) showed that, until 37 weeks gestational age, Asian babies grew in a remarkably similar fashion as those reported in the seminal Fenton charts but there was a significant and marked divergence after this. This may suggest the initiation of growth restriction at a consistent point in gestation in this cohort. We postulate that this restriction from the gestational age of 37 weeks may result from anatomical constraints due to the generally smaller Asian height, pelvic and uterine size. This postulation is supported by previous studies that indicated the shorter average gestational length by 1 week in Asian than Western pregnancies ^39,40^, and the shorter gestation associated with shorter maternal height ^33,41–43^. The shorter gestational length in Asian pregnancies is plausibly explained by the earlier maturation and senescence of the feto-placental unit in relation to maternal pelvic size ^33,41–43^ or a shorter cervical length ^44^. These findings may suggest that Asian normal gestational length might be shorter than that of European mothers as an evolutionary adaptation to smaller size. If it is indeed true that gestational length is race or ethnicity specific—a hypothesis that deserves further evaluation—we may need to change the definition of “term gestation” using ethnic-specific cut-off points. This proposal would potentially affect our clinical management, including the timing of delivery, classification of prematurity as well as perinatal management individualised to race or ethnicity. Separately our new birth anthropometry charts will impact clinical practice by more accurately defining normality in BW, length and head circumference. For example bedside glucose screening rates for at-risk infants will likely change as there would be fewer babies being labelled as small-for-gestational age (SGA)^45–47^. Surveillance for microcephaly was one key clinical characteristic used for detection of congenital Zika infection. Local HC charts would be needed for such a purpose^48–50^. Postnatal growth failure (PNGF), defined as a body weight below the 10^th^ percentile or a temporal weight loss of more than 1 or 2 standard deviation (SD) after birth, is seen commonly in infants born very preterm and affects their future neurodevelopment^51,52^. The use of growth charts appropriate for a particular population is thus useful for timely identification of PNGF and applying early nutritional intervention^53–55^.

### Strengths and Limitations

The strength of this study lies in the availability of a detailed and accurate database which allowed the examination of population data of three major Asian ethnicities. A comparatively uniform exposure to socioeconomic, cultural and healthcare influences of developed world standards allowed a level playing field for both inter-ethnic comparisons. This model provided a unique opportunity to unmask influences of genetic potential on fetal growth without being heavily confounded by external factors. A wide range of prenatal and perinatal determinants of fetal growth such as socio-economic status and maternal lifestyle factors (smoking, alcohol, and coffee intake) could be studied, variables which are sometimes not accurately captured in big population-based studies.

Our cohorts from 1991–97 and 2010-17 allow comparison to the original cohort used to develop the widely used Fenton chart (of 1991–2007, and revised in 2013 ^26^). Biological factors may not have changed much in the 2 decades that separate our 2 cohorts. However, SES, health care practices, management of high-risk pregnancy and maternal behavior have changed over the last two decades in Singapore. Some studies have demonstrated improving socioeconomic environment as well as increased maternal Body Mass Index (BMI), gestational weight gain, increased maternal height, less maternal smoking during pregnancy, and higher maternal education level to be responsible for progressive increase in BW ^56–60^. Contrary to this, several developed countries have reported a progressive decline in BW among the 21^st^ century babies^59–61^. This is likely multifactorial and possible causes could be attributable to changes in obstetric practices, increased proportion of pregnancies with maternal comorbidities and changes in maternal demographics resulting in earlier births and smaller babies^59–61^. The adjusted differences in BW, length and head circumference were small but significantly different between our 2 cohorts, with the newer cohort being slightly larger. We are currently comparing more detailed data between the 2 cohorts to unravel the underlying reasons.

## Conclusion

This study of birth anthropometry and its contributing factors among three Asian ethnic groups in Singapore showed that, contrary to published data, the three main maternal ethnicities among our population did not appear to be a strong predictor of birth anthropometry after controlling for other determinants like health, education and socioeconomic demographics. We also detected an apparent slowing of intrauterine growth velocity after 37 weeks gestation when comparing with international standards (Fenton), and as such the latter growth charts may not be appropriate for Singaporeans and perhaps Asians in general. We postulate that this observed apparent growth restriction in our cohort during the later weeks of pregnancy may be due to small Asian uterine and pelvic size, and early maturation and senescence of the feto-placental unit. Thus as a developmental adaptation, the normal Asian gestational length may well be slightly shorter than that in ethnic Europeans. Our data from a defined geopolitical area with stable racial and ethnic demography exposed to relatively uniform and high quality health, nutrition, socioeconomic factors forms an important baseline for future studies on developmental origins of health and diseases as well as for studying intergenerational trends. Our new growth charts allow more accurate determination of abnormality in birth size for the Asian population and has the potential to bring about better patient care practices.

## Data Availability

All data produced in the present study are available upon reasonable request to the authors

## Acknowledgement

This research is supported by the Singapore Ministry of Health’s National Medical Research Council under the Centre Grant Programme: Singapore Population Health Improvement Centre (NMRC/CG/C026/2017_NUHS).

## Disclosure of Interests

All authors declare no conflict of interests.

## Contribution to Authorship

LJ proposed the original concept and designed the study. LJ, SS, CA and ZA acquired the data. MYN and AC performed the statistical analysis. SS, AC, LJ, AB, ZA and MYN provided input on study design, analysed and interpreted the data. SS, AC and LJ drafted the manuscript. All authors approved the final version of the submitted manuscript.

## Details of Ethics Approval

The National Healthcare Group Domain Specific Review Board (DSRB) approved the study (DSRB Reference: 2018/00389 valid till 6 May 2022).

## Notes

The authors report no conflict of interest.

### Competing Interest Statement

The authors have declared no competing interest.

### Funding Statement

This study was funded by Singapore Population Health Improvement Centre (NMRC/CG/C026/2017_NUHS).

### Author Declarations

The National Healthcare Group Domain Specific Review Board (DSRB) gave ethical approval for the study (DSRB Reference: 2018/00389 valid till 6 May 2022).

## References

1. Rich-Edwards JW, Colditz GA, Stampfer MJ, et al. Birthweight and the risk for type 2 diabetes mellitus in adult women. Ann Intern Med. 1999;130(4 Pt 1):278–284.

2. Hovi P, Andersson S, Eriksson JG, et al. Glucose regulation in young adults with very low birth weight. N Engl J Med. 2007;356(20):2053–2063. doi:10.1056/NEJMoa067187

3. Oken E, Gillman MW. Fetal origins of obesity. Obes Res. 2003;11(4):496–506. doi:10.1038/oby.2003.69

4. Jeffery AN, Metcalf BS, Hosking J, Murphy MJ, Voss LD, Wilkin TJ. Little Evidence for Early Programming of Weight and Insulin Resistance for Contemporary Children: EarlyBird Diabetes Study Report 19. Pediatrics. 2006;118(3):1118–1123. doi:10.1542/peds.2006-0740

5. Gillman MW, Rifas-Shiman S, Berkey CS, Field AE, Colditz GA. Maternal gestational diabetes, birth weight, and adolescent obesity. Pediatrics. 2003;111(3):e221–226.

6. Rich-Edwards JW, Stampfer MJ, Manson JE, et al. Birth weight and risk of cardiovascular disease in a cohort of women followed up since 1976. BMJ. 1997;315(7105):396–400.

7. Sipola-Leppänen M, Karvonen R, Tikanmäki M, et al. Ambulatory blood pressure and its variability in adults born preterm. Hypertension. 2015;65(3):615–621. doi:10.1161/HYPERTENSIONAHA.114.04717

8. Loos RJ, Fagard R, Beunen G, Derom C, Vlietinck R. Birth weight and blood pressure in young adults: a prospective twin study. Circulation. 2001;104(14):1633–1638.

9. Ahlgren M, Wohlfahrt J, Olsen LW, Sørensen TIA, Melbye M. Birth weight and risk of cancer. Cancer. 2007;110(2):412–419. doi:10.1002/cncr.22773

10. Richards M, Hardy R, Kuh D, Wadsworth ME. Birth weight and cognitive function in the British 1946 birth cohort: longitudinal population based study. BMJ. 2001;322(7280):199–203.

11. Stein REK, Siegel MJ, Bauman LJ. Are children of moderately low birth weight at increased risk for poor health? A new look at an old question. Pediatrics. 2006;118(1):217–223. doi:10.1542/peds.2005-2836

12. McIntire DD, Bloom SL, Casey BM, Leveno KJ. Birth weight in relation to morbidity and mortality among newborn infants. N Engl J Med. 1999;340(16):1234–1238. doi:10.1056/NEJM199904223401603

13. Abel KM, Wicks S, Susser ES, et al. Birth weight, schizophrenia, and adult mental disorder: is risk confined to the smallest babies? Arch Gen Psychiatry. 2010;67(9):923–930. doi:10.1001/archgenpsychiatry.2010.100

14. Kelly A, Kevany J, de Onis M, Shah PM. A WHO Collaborative Study of Maternal Anthropometry and Pregnancy Outcomes. Int J Gynaecol Obstet. 1996;53(3):219–233.

15. Kleinman JC, Kessel SS. Racial differences in low birth weight. Trends and risk factors. N Engl J Med. 1987;317(12):749–753. doi:10.1056/NEJM198709173171207

16. Anderson JG, Rogers EE, Baer RJ, et al. Racial and Ethnic Disparities in Preterm Infant Mortality and Severe Morbidity: A Population-Based Study. Neonatology. 2018;113(1):44–54. doi:10.1159/000480536

17. Black RE, Allen LH, Bhutta ZA, et al. Maternal and child undernutrition: global and regional exposures and health consequences. Lancet. 2008;371(9608):243–260. doi:10.1016/S0140-6736(07)61690-0

18. Kramer MS, Séguin L, Lydon J, Goulet L. Socio-economic disparities in pregnancy outcome: why do the poor fare so poorly? Paediatr Perinat Epidemiol. 2000;14(3):194–210.

19. Brooke OG, Anderson HR, Bland JM, Peacock JL, Stewart CM. Effects on birth weight of smoking, alcohol, caffeine, socioeconomic factors, and psychosocial stress. BMJ. 1989;298(6676):795–801.

20. Wang X, Zuckerman B, Pearson C, et al. Maternal cigarette smoking, metabolic gene polymorphism, and infant birth weight. JAMA. 2002;287(2):195–202.

21. Mallia T, Grech A, Hili A, Calleja-Agius J, Pace NP. Genetic determinants of low birth weight. Minerva Ginecol. 2017;69(6):631–643. doi:10.23736/S0026-4784.17.04050-3

22. Lee WKM. The economic marginality of ethnic minorities: an analysis of ethnic income inequality in Singapore. Asian Ethn. 2004;5(1):27–41. doi:10.1080/1463136032000168880

23. Lim RBT, Zheng H, Yang Q, Cook AR, Chia KS, Lim WY. Ethnic and gender specific life expectancies of the Singapore population, 1965 to 2009 – converging, or diverging? BMC Public Health. 2013;13(1):1012. doi:10.1186/1471-2458-13-1012

24. GBD 2017 Disease and Injury Incidence and Prevalence Collaborators. Global, regional, and national incidence, prevalence, and years lived with disability for 354 diseases and injuries for 195 countries and territories, 1990-2017: a systematic analysis for the Global Burden of Disease Study 2017. Lancet. 2018;392(10159):1789–1858. doi:10.1016/S0140-6736(18)32279-7

25. Tan KL, Chia HP. Congenital Anomalies in Singapore. Congenit Anom. 1996;36(2):57–64. doi:10.1111/j.1741-4520.1996.tb00621.x

26. Fenton TR, Kim JH. A systematic review and meta-analysis to revise the Fenton growth chart for preterm infants. BMC Pediatr. 2013;13:59. doi:10.1186/1471-2431-13-59

27. Rigby RA, Stasinopoulos DM. Generalized additive models for location, scale and shape. :48.

28. R Core Development Team. R: A Language and Environment for Statistical Computing. R Foundation for Statistical Computing; 2019. https://www.R-project.org/

29. PediTools. Fenton 2013 Growth Calculator for Preterm Infants. https://peditools.org/fenton2013/

30. Chao F, Gerland P, Cook AR, Alkema L. Systematic assessment of the sex ratio at birth for all countries and estimation of national imbalances and regional reference levels. Proc Natl Acad Sci U S A. 2019;116(19):9303–9311. doi:10.1073/pnas.1812593116

31. David RJ, Collins JW. Differing Birth Weight among Infants of U.S.-Born Blacks, African-Born Blacks, and U.S.-Born Whites. N Engl J Med. 1997;337(17):1209–1214. doi:10.1056/NEJM199710233371706

32. Alexander GR, Kogan M, Bader D, Carlo W, Allen M, Mor J. US Birth Weight/Gestational Age-Specific Neonatal Mortality: 1995–1997 Rates for Whites, Hispanics, and Blacks. Pediatrics. 2003;111(1):e61–e66. doi:10.1542/peds.111.1.e61

33. Ozaltin E, Hill K, Subramanian SV. Association of maternal stature with offspring mortality, underweight, and stunting in low-to middle-income countries. JAMA. 2010;303(15):1507–1516. doi:10.1001/jama.2010.450

34. Kim SY, Sharma AJ, Sappenfield W, Wilson HG, Salihu HM. Association of maternal body mass index, excessive weight gain, and gestational diabetes mellitus with large-for-gestational-age births. Obstet Gynecol. 2014;123(4):737–744. doi:10.1097/AOG.0000000000000177

35. Shankaran S, Das A, Bauer CR, et al. Association between patterns of maternal substance use and infant birth weight, length, and head circumference. Pediatrics. 2004;114(2):e226–234.

36. Mills JL, Graubard BI, Harley EE, Rhoads GG, Berendes HW. Maternal alcohol consumption and birth weight. How much drinking during pregnancy is safe? JAMA. 1984;252(14):1875–1879.

37. Hinkle SN, Albert PS, Mendola P, et al. The association between parity and birthweight in a longitudinal consecutive pregnancy cohort. Paediatr Perinat Epidemiol. 2014;28(2):106–115. doi:10.1111/ppe.12099

38. World Bank. The World Bank Database. http://www.worldbank.org

39. Patel RR, Steer P, Doyle P, Little MP, Elliott P. Does gestation vary by ethnic group? A London-based study of over 122,000 pregnancies with spontaneous onset of labour. Int J Epidemiol. 2004;33(1):107–113. doi:10.1093/ije/dyg238

40. Savitz DA. Commentary: Ethnic differences in gestational age exist, but are they ‘normal’? Int J Epidemiol. 2004;33(1):114–115. doi:10.1093/ije/dyh041

41. Derraik J, Savage T, Hofman P, Cutfield W. Shorter mothers have shorter pregnancies. Int J Pediatr Endocrinol. 2015;2015(Suppl 1):P109. doi:10.1186/1687-9856-2015-S1-P109

42. Myklestad K, Vatten LJ, Magnussen EB, Salvesen KÅ, Romundstad PR. Do parental heights influence pregnancy length?: a population-based prospective study, HUNT 2. BMC Pregnancy Childbirth. 2013;13:33. doi:10.1186/1471-2393-13-33

43. Zhang G, Bacelis J, Lengyel C, et al. Assessing the Causal Relationship of Maternal Height on Birth Size and Gestational Age at Birth: A Mendelian Randomization Analysis. PLoS Med. 2015;12(8):e1001865. doi:10.1371/journal.pmed.1001865

44. Gagel CK, Rafael TJ, Berghella V. Is short stature associated with short cervical length? Am J Perinatol. 2010;27(9):691–695. doi:10.1055/s-0030-1253100

45. Mejri A, Dorval VG, Nuyt AM, Carceller A. Hypoglycemia in term newborns with a birth weight below the 10th percentile. Paediatr Child Health. 2010;15(5):271–275. doi:10.1093/pch/15.5.271

46. Adamkin DH, Committee on Fetus and Newborn. Postnatal Glucose Homeostasis in Late-Preterm and Term Infants. Pediatrics. 2011;127(3):575–579. doi:10.1542/peds.2010-3851

47. Abali S, Beken S, Albayrak E, et al. Neonatal Problems and Infancy Growth of Term SGA Infants: Does “SGA” Definition Need to Be Re-evaluated? Front Pediatr. 2021;9:660111. doi:10.3389/fped.2021.660111

48. Heukelbach J, Werneck GL. Surveillance of Zika virus infection and microcephaly in Brazil. The Lancet. 2016;388(10047):846–847. doi:10.1016/S0140-6736(16)30931-X

49. Ho ZJM, Hapuarachchi HC, Barkham T, et al. Outbreak of Zika virus infection in Singapore: an epidemiological, entomological, virological, and clinical analysis. Lancet Infect Dis. 2017;17(8):813–821. doi:10.1016/S1473-3099(17)30249-9

50. Harville EW, Tong VT, Gilboa SM, et al. Measurement of Head Circumference: Implications for Microcephaly Surveillance in Zika-Affected Areas. Trop Med Infect Dis. 2020;6(1):5. doi:10.3390/tropicalmed6010005

51. Latal-Hajnal B, von Siebenthal K, Kovari H, Bucher HU, Largo RH. Postnatal growth in VLBW infants: significant association with neurodevelopmental outcome. J Pediatr. 2003;143(2):163–170. doi:10.1067/S0022-3476(03)00243-9

52. Peila C, Spada E, Giuliani F, et al. Extrauterine Growth Restriction: Definitions and Predictability of Outcomes in a Cohort of Very Low Birth Weight Infants or Preterm Neonates. Nutrients. 2020;12(5):1224. doi:10.3390/nu12051224

53. Cole TJ, Statnikov Y, Santhakumaran S, Pan H, Modi N, on behalf of the Neonatal Data Analysis Unit and the Preterm Growth Investigator Group. Birth weight and longitudinal growth in infants born below 32 weeks’ gestation: a UK population study. Arch Dis Child - Fetal Neonatal Ed. 2014;99(1):F34–F40. doi:10.1136/archdischild-2012-303536

54. Andrews ET, Ashton JJ, Pearson F, Beattie RM, Johnson MJ. Early postnatal growth failure in preterm infants is not inevitable. Arch Dis Child - Fetal Neonatal Ed. 2019;104(3):F235–F241. doi:10.1136/archdischild-2018-315082

55. Lee LY, Lee J, Niduvaje K, Seah SS, Atmawidjaja RW, Cheah F. Nutritional therapies in the neonatal intensive care unit and post□natal growth outcomes of preterm very low birthweight Asian infants. J Paediatr Child Health. 2020;56(3):400–407. doi:10.1111/jpc.14634

56. Power C. National trends in birth weight: implications for future adult disease. BMJ. 1994;308(6939):1270–1271. doi:10.1136/bmj.308.6939.1270

57. Ananth CV, Wen SW. Trends in fetal growth among singleton gestations in the United States and Canada, 1985 through 1998. Semin Perinatol. 2002;26(4):260–267.

58. Fok TF, So HK, Wong E, et al. Updated gestational age specific birth weight, crown-heel length, and head circumference of Chinese newborns. Arch Dis Child Fetal Neonatal Ed. 2003;88(3):F229–236.

59. Donahue SMA, Kleinman KP, Gillman MW, Oken E. Trends in birth weight and gestational length among singleton term births in the United States: 1990-2005. Obstet Gynecol. 2010;115(2 Pt 1):357–364. doi:10.1097/AOG.0b013e3181cbd5f5

60. Oken E. Secular trends in birthweight. Nestle Nutr Inst Workshop Ser. 2013;71:103–114. doi:10.1159/000342576

61. Catov JM, Lee M, Roberts JM, Xu J, Simhan HN. Race Disparities and Decreasing Birth Weight: Are All Babies Getting Smaller? Am J Epidemiol. 2016;183(1):15–23. doi:10.1093/aje/kwv194

